# Containment to outbreak tipping points in COVID-19

**DOI:** 10.1101/2020.09.14.20194159

**Authors:** Matías Arim, Daniel Herrera-Esposito, Paola Bermolen, Álvaro Cabana, María Inés Fariello, Mauricio Lima, Hector Romero

## Abstract

Non-pharmaceutical interventions (NPIs) have been a cornerstone in managing emergent diseases such as COVID-19^1–4^. However, despite their potential to contain or attenuate the epidemic, the effects of NPIs on disease dynamics are not well understood^1,5–7^. We show that saturation of NPIs with the increase in infected individuals, an expected consequence of limited contact tracing and healthcare capacities, produces a positive feedback in the disease growth rate and a threshold between two alternative states--containment and outbreak^8^. These alternative states were previously related with the strength of NPIs but not with the infection number^2,9–11^. Furthermore, the transition between these states involves an abrupt acceleration in disease dynamics, which we report here for several COVID-19 outbreaks around the world. The consequences of a positive feedback in population dynamics at low numbers is a phenomenon widely studied in ecology--the Allee effect. This effect is a determinant of extinction-outbreak states, geographic synchronization, spatial spread, and the effect of exogenous variables, as vaccination^12–15^. As countries are relaxing containing measures, recognizing an NPI-induced Allee effect may be essential for deploying containment strategies within and among countries^16^ and acknowledges the need for early warning indicators of approaching epidemic tipping points^17^.

---

Non-pharmaceutical interventions (NPIs) have been applied with unprecedented strength and breadth in managing the COVID-19 world pandemic^2–4,7^. These interventions involve case isolation, contact tracing, testing and quarantine, reduction in the number of contacts by social distancing, and behavioural changes oriented to reduce transmission—e.g., mask use and spatial distancing^2,10,18^. As a general rule, both observed dynamics and model projections indicate that the implementation of strong interventions significantly reduced the total number of infections compared with uncontained outbreaks^2,3,6,7,19,20^. The effectiveness of NPIs may consequently determine two alternative states: one of disease containment (*R_e_<1*) and another of disease outbreak (*R_e_>1*)^2,9–11^. However, these two states were observed among regions with similar strengths of NPIs and through time in countries, highlighting the need for a better understanding of the mechanisms guiding the impact of NPIs in COVID-19 dynamics^1,4–7^.

An intrinsic characteristic of NPIs is that their effectiveness is expected to wane as the number of infected individuals increases^10^. For example, the larger the number of infections, the lower the probability of performing a complete tracking of all the contacts of an infected individual^10^. Similarly, the effect of face masks, hygiene measures, and physical distance on the transmission rate is expected to decrease with a higher pathogen load in the environment^18^. Finally, the healthcare system’s progressive occupation may extend the infectious period, producing more social interactions when looking for medical attention, delays in case isolation, and low availability of protective material^21,22^.

This saturation of NPIs generates a positive feedback between the number of infected individuals and disease growth rate, which has critical consequences in the disease dynamics, particularly at low numbers. This feedback loop connects the saturation of NPIs with the Allee effect, a central concept in population biology that explains the fate of several populations at low numbers^12,13^. Indeed, the Allee effect involves a positive feedback between population abundance and population growth rate, which may determine a transition from a negative to a positive growth rate at a given abundance threshold^12,13^. In disease dynamics, this is interpreted as an epidemic breakpoint, a tipping point below which the outbreak tends to diminish (*R_e_<1*), but above which the outbreak grows (*R_e_>1*)^8^. However, this phenomenon is not captured in the many variants of SIR and logistic models widely used for the analysis and forecasting of COVID-19 dynamics^2,3,6,7,19,23^. Here we aim to fill this gap, formalizing the connection between saturating NPIs and disease dynamics. We show how the saturation of NPIs induce a positive feedback in disease dynamics, from which abrupt transitions in spread rate are expected after a threshold in the number of infected individuals is crossed. Finally, we look for traces of this footprint in actual data from the COVID-19 pandemic.

## Allee effect induced by NPIs

In the absence of contact tracing and quarantine, a randomly chosen infected individual will produce an average number of second infections *R_e^nq^_* (*nq*: non quarantine). For the sake of clarity in our model we will use quarantine to encapsulate the concepts of ‘quarantine’ and ‘case isolation’ without losing explanatory power. This value is determined by the number of social links accumulated in the infectious time (*L_max_*), the probability of disease transmission in each link (*b_link_*), and the probability of having a link with a susceptible individual (*P_susceptible_*) thus *R_e_^nq^* = *P_susceptible_*·*b_link_*·*L_max_* (Fig. 1A).

**Figure 1.**
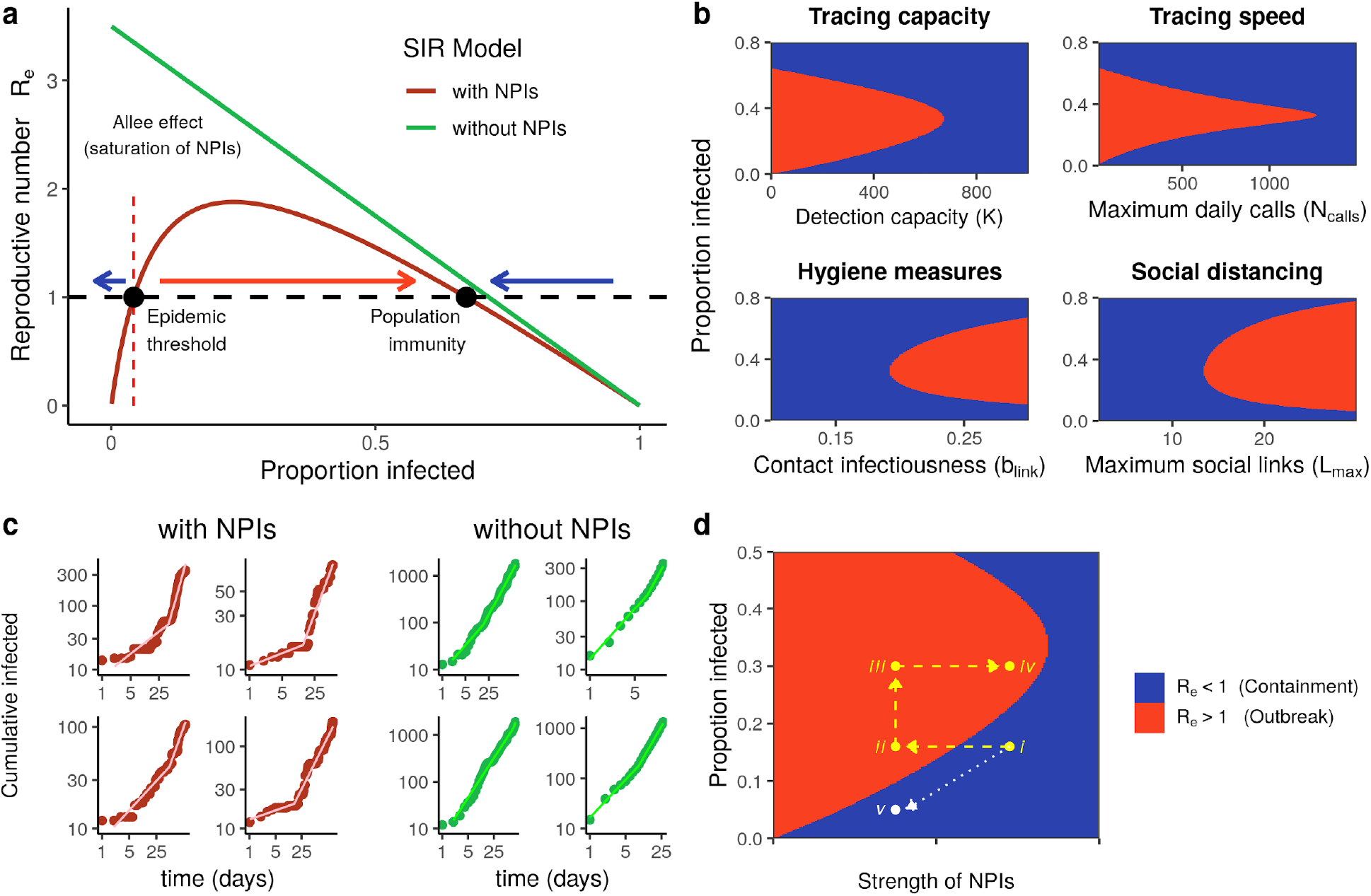
Saturation of NPIs and disease dynamics. **(a)** Effective reproduction number as a function of the proportion of the population infected *R_e_(I)*. Without NPIs (green line) the per capita growth rate *R_e_(I)* decreases proportionally to the infection number indicating a logistic dynamic^13^. When NPIs are in place (red line) *R_e_(I)* first increases with the proportion of infected individuals due to the saturation of NPIs and then decreases approaching the dynamics in absence of NPIs (equation [1]). This determines a positive feedback at low numbers (NPI-Allee effect) and a threshold for epidemic outbreak in the number of infections (*I\**). (**b**) The alternative epidemic states of growth (*R_e_(I)>1*, red) or containment (*R_e_(I)<1*, blue) are determined by both the proportion of the infected population (vertical axis) and the strength of the different NPIs (horizontal axis) **(c)** SIR dynamics without NPIs (right) and with NPIs of limited capacity (left), inducing an Allee effect that can generate abrupt transitions in disease spread rate (equation [2]). **(d)** Relaxation of NPIs and epidemic containment. In trajectory *i*→*ii*→*iii*→*iv* an abrupt relaxation surpasses the epidemic threshold provoking an increase in infections. Return to previous strength of NPIs fails to contain the outbreak. In trajectory *i*→*v* the gradual relaxation of NPIs follows the decrease in the infected population keeping the epidemic under control.

Strong enough NPIs will determine a *R_e_* <1 controlling the epidemic^2,9–11^. While face mask use, hygienic measures, and physical distancing reduce *b_link_*^18^, the number of social links *L_max_* can be reduced through contact tracing and subsequent quarantine^10^. Individuals detected by the tracing system are contacted and then put on quarantine. However, only a fraction *f_q_* of the infected individuals is detected (*D = f_q_·I*) by the contact tracing, which is determined by the maximum number of cases (*K*) that can be processed in a day. Moreover, because of the saturation of tracing systems, as the number of infected individuals increases, the fraction *f_q_* decreases—here modeled as *f_q_ = K/(I_50,D_+I)*, with *I_50,D_* being the number of infected cases at which half the maximum detection is reached^24^.

The saturation of the contact tracing system increases the time lag between infection and detection. This is modeled considering that the tracing system can make a fixed number of calls (*N_calls_*) each day and that each contact attempt has a probability of success *P_find_*. Consequently, the expected number of alerts received per individual on a given day is *N_calls_/D*, and the probability of finding and quarantining him is *P_q_ = 1-(1-P_find_)^Ncalls/D^*. The probability that an individual is contacted and quarantined on day *d* after the infection onset follows the geometric distribution *P(d) = P_q_(1-P_q_)^d-1^*. We assume that during this period, individuals establish social links with infectious potential^25^ according to the function *L(d) = L_max_·d^4^/(d^4^+7^4^)* (see Supplementary Methods). Then, the expected number of links generated before quarantine is: *L = Σ_d≥1_ P(d)·L(d)*.

Integrating all of the above, we can express the effective reproduction number *R_e_(I)* determined by the second infections generated by both fractions of quarantined *f_q_* and non quarantined *f_nq_* infected individuals as:

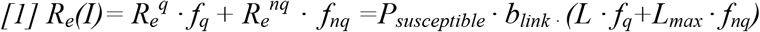

This equation’s plot in Figure 1a shows that NPIs induce a positive relationship between the infected individuals in a population and the epidemic reproduction number. In particular, it is expected that the epidemic does not take-off at low infection numbers (*R_e_(I)<1*), requiring a minimum level of infections (*I\**) to cause an outbreak (*R_e_(I)>1*).

The strength of the different components of NPIs—contact tracing capacity and speed, social distance, and transmission barriers— have nonlinear relationships with the outbreak threshold *I\** (Fig. 1b). Alternative states^8,17^ of growing and diminishing epidemic was considered elsewhere for COVID-19 but as a response to the strength of NPIs^2,9–11^. Here, we show that for a fixed level of saturating NPIs, the transition from contention to outbreak (i.e. *R_e_(I)<1*⇒ *R_e_(I)>1*) may be determined by the number of infected individuals (Fig. 1a, b).

## Disease dynamics with NPI induced Allee effect

In a deterministic system with a saturation of NPIs that induces an Allee effect (hereafter NPI-Allee effect), infections are expected to either vanish or take-off. However, at low numbers, pulses of immigration of infected individuals and super spreading events could determine the transition from a negative to a positive disease growth rate if the threshold is exceeded^26^. In this section, we assess the qualitative consequences of the saturation of NPIs on disease dynamics through a SIR model considering stochasticity and infection pulses with a heavy tailed distribution.

The NPI-Allee effect is introduced in the daily rate of disease transmission through a saturating function *β_t_(I) = β_max_·I(t)/(I_50.β_+I(t))* and also in the average duration of the infection period, *γ_t_(I) = γ_max_·I(t)/(I_50.γ_+I(t))*:

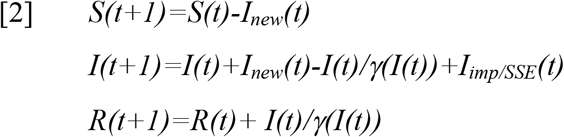

Where *I_new_(t)∼Poisson(λ(t))* with *λ(t) = βt(I)·I^p^(t)S(t)/N*; the exponent *p* accounts for the prevalence of subexponential dynamics in Covid-19 related to the effect of NPIs^7^;*I_imp/SSE_(t)∼NB(m,s)* is a negative binomial distribution^27^ with parameters *m* (mean) and size *s* (dispersion) and refers to the importation of new cases and super spread events (SSE, see Supplementary Methods). Trajectories of this model were simulated to observe the impact of the NPI-Allee effect on the evolution of the cumulative number of infected individuals (*C_t_*). We compared these trajectories to those obtained with the model without NPI saturation, with fixed *β* = *β_max_* and, *γ = γ_max_* (Fig. 1c and Extended Data Figure 1).

The logarithm of the cumulative number of infections grows linearly in time in exponential dynamics^13,24^. However, as observed in COVID-19, sub-exponential dynamics grow linearly with the logarithm of time^7^, as described by the relationship *log(Ct) = µ·log(t)*. We use this time scale to visualize our simulations, where the parameter *µ* describes the disease dynamics. The SIR model without the NPI-Allee effect presents a consistent trend with minor changes fueled by the stochastic effects of imported cases and super spread events (Fig. 1c, Extended Data Figs. 1). On the other hand, when the NPI-Allee effect is considered, we observe a qualitative transition in the dynamics: from a slow rate of case accumulation to a markedly larger one, suggesting that these random events may determine the surpass of a tipping point from containment to outbreak. We fitted a segmented regression to examine the differences in dynamics with and without NPI-Allee effect (Fig. 1c, Extended Data Fig 1). Both simulations present breaking points and may look similar when this point is rapidly reached. In order to further explore their difference, we plotted the relative increment of the slope after the breaking point (*µ2/ µ1*) as a function of the initial slope(*µ1*) for both kinds of simulations (Fig. 2a and 2c). Now, it becomes apparent that including saturating NPIs expands the range of observed dynamics, promoting slower rates during the initial phase and, after the tipping point, an accelerated dynamic capturing the loss in effectiveness of the NPIs (Figs. 2a and 2b, Extended Data Fig. 1). Our model results show how NPIs saturation may induce an Allee effect that produces abrupt transitions, with periods of disease containment and periods of rapid growth. In this sense, the role of a heavy-tailed distribution in new infections markedly differs between the two models, because super spread events, and/or imported cases, may cause the infection number to exceed the threshold.

**Figure 2.**
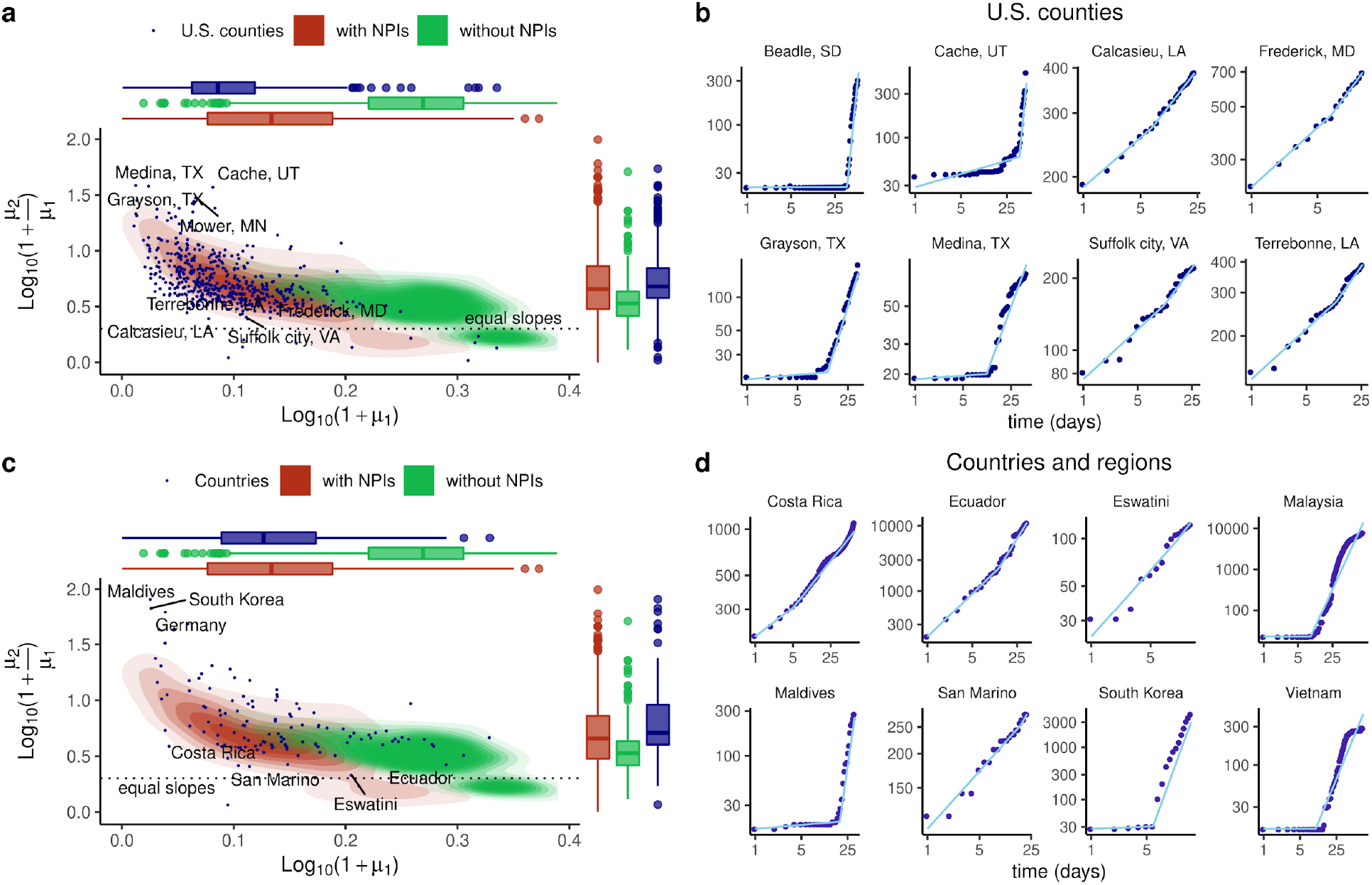
Observed dynamics of cumulative cases of COVID-19 and SIR models simulations with and without NPI-Allee effect. Trajectories are described by segmented regressions on *log(Ct) = µ·log(t)* with the slopes *µ1* and *µ2* representing the rate of cases accumulation before and after the breakpoint. **(a)** COVID-19 dynamics in U.S. counties (blue circles) frequently shown an initial phase with low rate of increase in cumulative cases followed by a transition to a faster growth rate. The initial slope (*µ1*) is shown in the horizontal axis as *log_10_(1+ µ1)* and the relative increment in slope after the breaking point (*µ2/ µ1*) is shown in the vertical axis as *log_10_(1+ µ2/ µ1)*. This dynamic is frequently observed in the SIR model with NPI-Allee effect (red contours) but not in models without this effect (green contours). Red and green density plots were estimated from 1000 simulated SIR dynamics with and without NPI-Allee effect, with parameters: *β*_max_ = 0.5, *γ*_max_ = 4, p = 0.8, m = 1, I_50.γ_ = I_50.β_ = 10 and *s* = 0.2, with population sizes sampled from a log-Normal distribution with equal mean and variance than U.S. counties populations. Simulations covering a wide range of plausible parameters for COVID-19 consistently support the congruence of observed dynamics with the NPI-Allee effect model (Supplementary Methods and Extended Data Fig. 5). **(b)** Range of dynamics observed for U.S. Counties. Segmented regressions always provide a close description of the dynamics of cumulative cases. **(c and d)** same analysis than **(a)** and **(b)** but for Countries and regions.

## Tipping points in actual COVID-19 data

Guided by the previous results, we looked for the existence of tipping points and acceleration of case accumulation of COVID-19 dynamics in 532 U.S counties and 125 countries. As we focus on the initial dynamics, we consider the time frame starting 14 days after surpassing 10 accumulated cases, and finishing the day with the maximum number of daily infections. Based again on the relationship *log(Ct) = µ·log(t)*, we fitted a segmented regression and contrasted its performance with a linear fit without a breaking point. Almost all of the observed dynamics were better described by a segmented relationship, suggesting that breaking points are a common phenomenon in COVID-19 dynamics (Fig. 2; Extended Data Figs. 2, 3, and 4). Further, the segmented model provides an excellent description of the dynamics of cumulative cases of Covid-19, with an average R^2^ of 0.99 across U.S. counties and countries (Extended Data Fig. 4).

The observed dynamics show several cases with a striking similarity to the NPI-Allee effect simulations, with abrupt transitions from slow to fast growth rate and a delayed tipping point. When we project the US-county data and worldwide country/region data of COVID-19, the results are eloquent (Fig. 2b and 2d and Extended Data Figs. 2 and 3). Indeed, the area covered with actual data is remarkably similar to the simulated cases with NPI-Allee effect. On the other hand, many regions do not show this behaviour, either because the breaking point occurred early in the epidemic spread-e.g., surpassing *I\** before NPIs were fully implemented--or because the NPIs have not determined the *I\** threshold in these locations.

## Discussion

Amidst an ongoing worldwide outbreak of COVID-19, studies connecting NPIs and disease dynamics are all the more needed^1,4–7,16^. Our results link NPIs with the substantial knowledge built around the Allee effect and tipping points in ecology^12,15,17,26,28^, and previous studies that related the saturation of NPIs, reactive individual behaviour, and positive feedbacks with disease dynamics^8,21,22,29^. The recognition of an NPI-Allee effect and the existence of tipping points contributes to the understanding of several features of disease dynamics^12,13,26^.

An immediate consequence is the understanding that the timing of NPIs is crucial. Interventions implemented when the disease has had enough time to surpass the correspondent threshold will fail in its containment^8,10,22^. Similarly, the relaxation of NPIs has to be done in attention to the epidemic threshold for avoiding outbreak resurgence. The idea of relaxing interventions and then returning them to former levels if the number of infected individuals increases may be futile after the tipping point. NPIs may be relaxed cautiously following the decrease in the number of infections with attention to potential tipping points (Fig. 1d), a context where early warning indicators become crucial. These indicators were related to systems resilience and increased trajectory variance^17^, but more specific indicators for diseases under NPIs management are probably required. For example, in a contained state of the disease, the fraction of reported infections with no epidemiological link (not detected by the tracing system), and the time between individual exposure and quarantine should decrease through time. A systematic increase in these indicators may be an adequate early warning signal that containment could be compromised.

Another important feature is that NPIs could operate as exogenous variables synchronizing disease dynamics along large geographic scales^28,30^, being more likely to simultaneously control the outbreaks or to enable global resurgence^14,15^. The success of proposed spatial synchrony of NPIs for controlling COVID-19^16^ and vaccination strategies may depend on the associated NPI-Allee effect’s strength. More generally, the spatial spread of diseases with NPI-Allee effect may be guided by local surpass of epidemic threshold and neighbor contagion^15,26^. In diseases around an outbreak tipping point, spatial and temporal mosaic of *R_e_>;1* and *R_e_<1* may arise, determining local outbreaks, and fostering disease persistence. Finally, we showed how the dynamic consequences of super spread events are contingent to its effect on saturating NPIs.

Recognizing the NPIs-induced Allee effect has large consequences for the understanding and management of COVID-19 and other diseases and may significantly expand the theoretical framework for the application of NPIs in ongoing and future epidemics. This knowledge is of particular relevance for explaining the past dynamics of COVID-19 in different regions of the world and, more importantly, as input for guiding NPIs relaxation strategies and preventing new outbreaks.

## Data Availability

Data Availability
Country and regional COVID-19 daily data and demographic data were obtained from Johns Hopkins University COVID-19 Data Repository31.
Code Availability
All the code used in this work is available at https://github.com/almadana/alle.covid/.

https://github.com/almadana/alle.covid/

## Data Availability

Country and regional COVID-19 daily data and demographic data were obtained from Johns Hopkins University COVID-19 Data Repository^31^.

## Code Availability

All the code used in this work is available at https://github.com/almadana/alle.covid/.

## Acknowledgements

Authors thank comments from GUIAD and GACH. MA. thanks CSIC groups-657725. This work was partially supported by CSIC-UdelaR, PEDECIBA and ANII.

## Author Contributions

MA, HR designed the study; DH, AC, MA wrote computer codes and performed the simulations; DH, AC, MIF, HR, MA performed analysis of COVID-19 dynamcis; MA, HR wrote the first draft of ms; PB wrote the first draft of Supplementary Methods and models description. All authors discussed and contributed to models formulation and actively participated in writing the ms. ML contributes in developing the idea of NPI-Allee effect.

Supplementary Information is available for this paper.

Correspondence and requests for materials should be addressed to DH and AC.

